# Optical Coherence Tomography-Angiography Metrics in Patients with Atrial Fibrillation and Cerebral Microbleeds

**DOI:** 10.1101/2023.09.03.23294999

**Authors:** Yannie Soo, Carol Y. Cheung, Dawei Yang, Jill Abrigo, Bonnie Lam, Huijing Zheng, Suk Fung Tsang, Bonaventure Ip, Winnie Chu, Vincent Mok, Thomas Leung, the iSAVE Investigators

## Abstract

**Background:** Presence of cerebral microbleeds (CMB) on MRI brain, increases risk of intracerebral haemorrhage in patients with atrial fibrillation (AF) who require anticoagulation. The retina shares similar embryological and pathological properties with cerebral vessels. Studying changes in retinal vasculature at capillary level associated with presence of CMB may provide complementary prognostic information to assess risk of anticoagulant-related intracerebral hemorrhage.

We aimed to investigate changes in the retinal capillary network associated with presence of CMB in patients with AF using Optical Coherence Tomography-Angiography (OCT-A).

**Methods:** Patients with AF were prospectively recruited to undergo both OCT-A and MRI brain. OCT-A metrics of the superficial capillary plexus (SCP) and deep capillary plexus (DCP) at macular region, along with the radial peripapillary capillary network at disc center were measured quantitatively by an automated image analysis program. Multivariable logistic regression and quasi-poisson regression were performed to investigate the association of OCT-A metrics with presence and burden of CMBs respectively. Significant OCT-A metrics were compared between patients with AF and healthy control subjects.

**Results:** Among 99 patients with AF, 29 patients had CMB. A larger foveal avascular zone (FAZ) in SCP, also higher vessel diameter index (VDI) across SCP, DCP and disc center were significantly associated with presence and burden of CMB, after adjusting for age, diabetes mellitus, white matter hyperintensities ratio, image quality index, age, duration between OCT-A and MRI. A lower vessel density in SCP was associated with presence but not burden of CMB. Compared with healthy control, FAZ and VDI across SCP, DCP and disc area remained consistently different between patients with AF and control group.

**Conclusions:** This pilot study provides preliminary data supporting potential role of OCT-A to assist risk-stratification of bleeding-prone microangiopathy for patients with AF who require anticoagulation. Further studies with larger sample size and different ethnic groups are needed.

## Introduction

Atrial fibrillation (AF) accounts for one-third of the ischaemic stroke has been on growing trend with the aging population.(1–3) Despite the effectiveness of oral anticoagulants which reduces stroke risk by almost 60-70%, the possible risk of life-threatening intracerebral haemorrhage remains a common clinical challenge in treatment decisions. This is of particular concern in patients who have multiple cerebral microbleeds (CMB) on MRI brain. CMB are dot-like hypointense signals on MRI Susceptibility-weighted or Gradient Echo Imaging. They are hemosiderin deposits in macrophages after silent perivascular leakage, a biomarker indicating underlying bleeding-prone microangiopathy.

Data from meta-analysis have shown that patients with AF have increased risk of anticoagulant-associated intracerebral haemorrhage, which increases with burden of CMB (OR 2.68 for patients with CMB and OR 5.5 for patients with ≥5 CMB). (4) The frequent presence of CMB in around one-third of patients with AF has aroused research interests to integrate CMB evaluation in the algorithm for anticoagulation decisions. In the recent MICON study in patients with AF, patients with multiple CMB receiving warfarin had subsequent incident rate of intracerebral haemorrhage comparable to that of ischaemic stroke. While patients on combination of anticoagulant and antiplatelet had incident rate of intracerebral haemorrhage higher than that of ischemic stroke. (5)

Although MRI brain provides informative prognostic information, there are many limitations. First, it is a costly examination with limited access. Hence, it is seldom performed in patients with AF who have not had stroke or transient ischaemic attacks that these patients are often excluded in studies in this field. Furthermore, many patients with AF have relative contraindications for MRI (e.g. cognitive impairment, dysphasia or severe disability after stroke, presence of pacemakers, metallic heart valves etc), as reflected by the 20% screening failure rate in the IPAAC (Risk of intracerebral haemorrhage in Patients taking oral

Anticoagulants for Atrial fibrillation with Cerebral microbleeds) study.(6) Developing a non-invasive method for evaluating bleeding-prone microangiopathy for patients with AF at a lower cost, which can be performed in patients with and without history of stroke or transient ischaemic attack, may improve their accessibility to risk-stratification tools in the community which help individualise treatment decisions.

Being the third and inner coat of the eye, the retina shares similar embryological origin, anatomical features with the brain.(7, 8) Hence, many pathological changes in cerebral vessels are also reflected by alterations in the retinal vessels. Histopathological studies have shown that subjects with stroke had similar pathological changes such as fibrinoid degeneration and fibrohyalinoid thickening observed both in retinal arterioles and the brain. (9) Optical Coherence Tomography-Angiography (OCT-A) is a novel non-invasive technique for imaging microvasculature of the retina and choroid. Based on mapping erythrocyte movement over time by comparing sequential OCT B-scans (motion contrast) at a given cross-section, OCT-A provides 3-dimensional visualization of the capillary networks in the eye without need of intravenous dye injection.(10) With the semi-automated computer software, vascular parameters in the different layers in the retina, including vessel caliber, branching pattern, vascular density and the foveal avascular zone (i.e. the OCT-A metrics) can be quantified meticulously for analysis.

### Aim and Objectives

The aim of this study was to identify changes in the retinal capillary network associated with presence of CMB on MRI brain in patients with AF using OCT-A. We hypothesized that patients with CMB were more likely to have alterations in OCT-A metrics compared to those without CMB.

## Methods

### Patient Selection

In this prospective study, patients were recruited from Medical out-patient clinics at Prince of Wales Hospital during the period from November 2018 to Jan 2022. Inclusion criteria were Chinese patients aged ≥ 18 years; evidence of atrial fibrillation or atrial flutter documented on electrocardiogram, or holter. Exclusion criteria included contraindications for MRI brain (e.g. uncooperative patients, presence of pacemaker or metallic heart valve etc.), poor sitting balance to carry out retinal photography, known intracranial and ocular pathologies or previous surgery which may affect retinal image or MRI scan quality (e.g. brain tumour, dense cataract, corneal ulcer, glaucoma, retinal detachment etc) and pregnant patients. Informed consents were obtained. MRI brain were performed at Prince of Wales Hospital, while OCT-A were performed within 2 weeks before or after MRI brain at Hong Kong Eye Hospital. The study has been approved by regional research ethics committee. Data were presented according to the STROBE guidelines. Anonymized data of this study will be available from Corresponding Author upon justified requests.

A sample size of 135 patients was planned with the assumptions of (i) prevalence of CMB was 35% in patients with AF, (ii) 13% of patients with CMB had ≥5 CMB and (iii) a dropout rate of 10%.

To examine the occurrence of retinal vascular parameters associated with CMB in normal subjects, 65 healthy subjects aged ≥ 60 years, with no known ophthalmological or retinal diseases, AF, cardiovascular diseases, history of ischaemic or haemorrhagic stroke, diabetes mellitus and hypertension were recruited from the community for retinal imaging to serve as the control group.

Furthermore, to explore if changes in OCT-A metrics were also associated with longitudinal progression of CMB count, a subgroup of patients who had prior MRI performed during their enrolment in the IPAAC registry was identified.(11) The CMB count in the two sets of MRI were compared for development of new CMB. OCT-A metrics were compared between patients with and without new CMB in the follow-up MRI (i.e. index MRI for this study).

### Imaging of Capillary Networks Using OCT-A

All recruited subjects had their pupils of a randomly selected eye dilated with tropicamide and phenylephrine (0.5% each) and underwent OCT-A with Triton swept-source OCT (DRI-OCT, Topcon, Inc, Tokyo, Japan), which contains a swept-source with a light source of 1,050nm wavelength and a speed of 100,000 A-scans per second. Volumetric OCT scans centered on the fovea and optic disc were obtained for a scan area of 3×3 mm containing 320×320 A-scans. We used the built-in software (IMAGEnet6, v1.23.15008, Basic License 10) to generate OCT-A of superficial capillary plexus (SCP) and deep capillary plexus (DCP) at macular region and the radial peripapillary capillary network at disc center region, which gave improved detection sensitivity for low blood flow and reduced motion artefacts.(12) (**Figure 1**)

**Figure 1.**
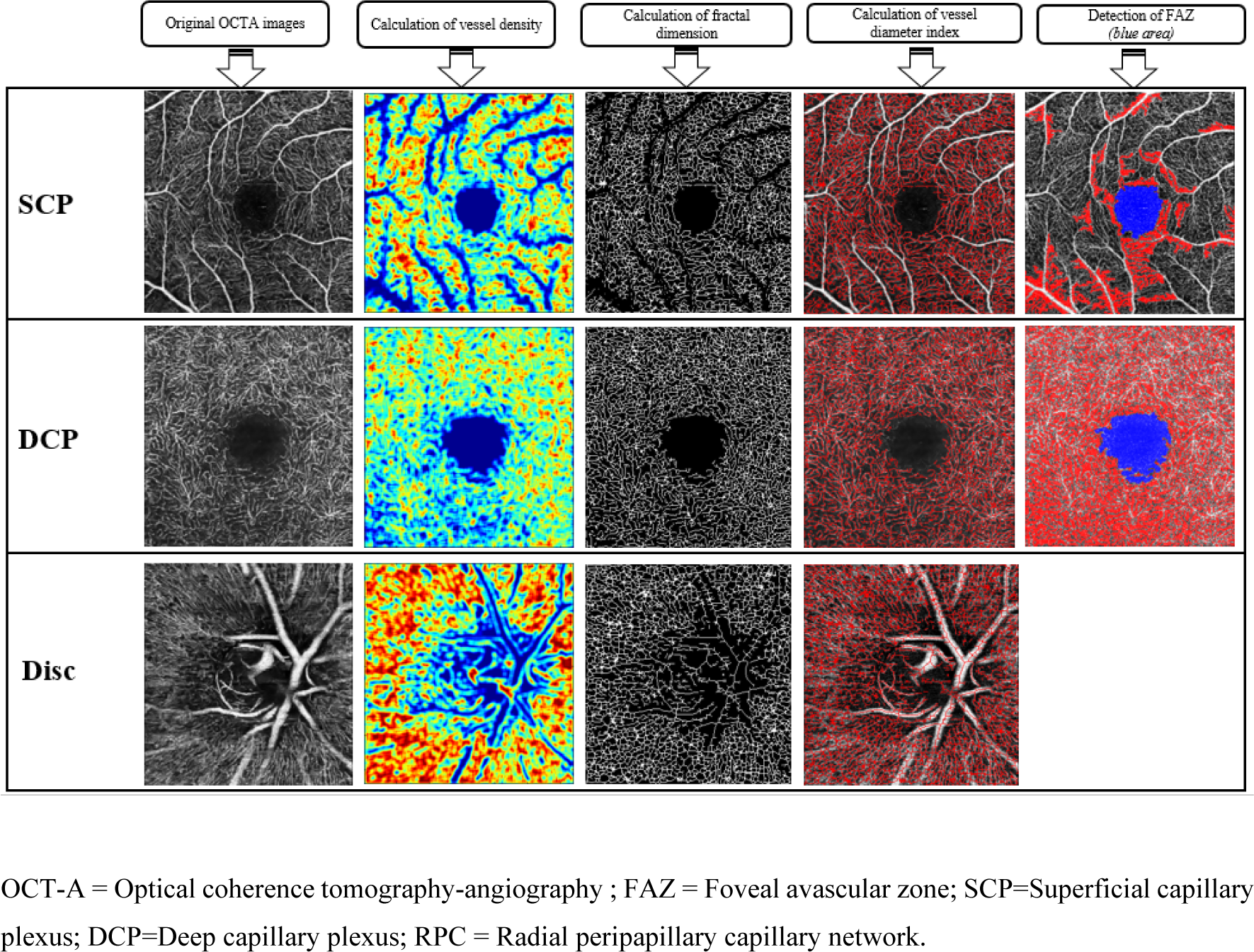
A spectrum of OCT-A metrics are measured on the OCT-A images of superficial capillary plexus, deep capillary plexus and radial peripapillary capillary network at disc center.

### OCT-A Images Quality Control

All OCT-A and OCT cross-sectional B-scan images were carefully evaluated in the CUHK Ocular Reading Centre before the quantitative analysis. Image graders were masked to all patient characteristics. An ungradable OCT-A image was defined as any artifacts concerning the FAZ area and/or impeding the identification of microvasculature or non-perfusion area.(13)

### Quantification of Capillary Network

OCT-A images were exported in grayscale from the built-in software. The images were then imported into an automated customized MATLAB program for image analysis.(12, 14) The area of the fovea avascular zone (FAZ) (mm^2^) in SCP and DCP was respectively calculated by counting total numbers of pixels within the region in scale. Vessel density (VD) (%) was defined as the percentage of area not defined as non-perfusion regions over total area within the interested region, of which in the macular area it was calculated over the Early Treatment Diabetic Retinopathy Study grid (1, 3mm), while in the disc area it was calculated over the circumpapillary region. (15)(16) Vessel diameter index (VDI), which reflects the vessel caliber, was calculated as the area occupied by blood vessel from the binarized image over the total length of blood vessel from the skeletonized image.(16).Fractal dimension, which represents a global measurement of the complexity of the vascular branching pattern, was also automatically determined by the MATLAB program from the skeletonized image using the box-counting method.(17, 18) (Figure 1)

### Neuroimaging

MR examinations were performed on a 3T scanner (Achieva TX; Philips Medical Systems, Best, Netherlands) using an 8-channel head coil. All patients underwent our standard stroke scanning protocol. Venous BOLD parameters were TR/TE=18/25ms, flip angle=15° and reconstructed at 0.45×0.45×1.0mm. FLAIR parameters were TR/TI/TE=11000ms/2800ms/125ms, flip angle=90°, reconstructed at 0.33×0.33×5.0mm.

CMB were rated by neuroradiologist (JMA) and neurologist experienced in rating CMB (YS), who had inter-rater reliability of 0.82 (p<0.001). A CMB was defined as an old, silent focus of signal loss in the Venous BOLD imaging, measuring 2-10 mm in diameter. CMB were distinguished from their mimics (e.g. basal ganglia calcifications, flow void artefacts of pial blood vessels etc) by correlating their morphologies on T1-, T2-weighted and diffusion weighted images. White matter hyperintensities on FLAIR sequence were scored using a visual rating scale proposed by Fazekas et al. (19) In addition, severity of white matter hyperintensities relative to the brain volume was expressed as the ratio of volume of white matter hyperintensities / brain parenchyma volume, which was evaluated by automated segmentation computational method. (20) Patients with movement artefact degrading quantification of CMB, white matter, brain volume or their segmentation were excluded.

### Statistical Analyses

Baseline characteristics and retinal parameters were compared between patients with and without CMB. Chi-square or Fisher exact test was used for categorical variables, while independent t-test or Mann-Whitney U test was used for continuous variables after normality examination. Same methods were used to compare retinal parameters between AF and control group. Continuous variables were expressed as mean ± standard deviation (SD) or median (interquartile range [IQR]).

Multivariable logistic regression models were applied to examine the associations of OCT-A metrics with presence of CMB, while the associations of with CMB count was examined with quasi-poisson regression models. All models were adjusted for age, white matter hyperintensities volume ratio, duration between OCT-A and MRI scans, diabetes mellitus and OCT-A image quality index. In the subgroup of patients who had prior MRI brain, OCT-A metrics were compared between patients with and without new CMB on follow-up MRI (i.e. index scan) using independent t-test or Mann-Whitney U test.

A p value < 0.05 was considered statistically significant. All statistical analyses were performed using R Statistical Software (version 4.3.0)

## Results

A total of 135 patients with AF and 65 control subjects were recruited. Due to the COVID-19 pandemic, 13 subjects withdrew from the study. After excluding 20 subjects with suboptimal OCT-A image quality, 1 subject with suboptimal MRI image quality and 1 subject with suboptimal image quality with both OCT-A and MRI, 99 patients with AF and 60 control subjects were included in the final analysis.

The mean age of the patients with AF was 71±11.5 years, 52.5% were male. The median CHA2D-VAS2C score was 4 (IQR = 3,5), while HAS-BLED score was 2 (IQR = 2,3). All patients were on direct oral anticoagulants, none were on Vitamin K antagonist nor antiplatelet. Due to delays in imaging during the COVID-19 pandemic, the median duration between retinal image and MRI scan was 12 days (IQR = 6, 29.5).

### Characteristics of Patients with and without CMB

CMB were identified in 29 (29.3%) patients. Among patients with CMB, the median CMB count was 2 (IQR 1,3). Compared to patients without CMB, patients with CMB had significantly higher white matter hyperintensity. There was otherwise no significant difference in demographic, clinical background and duration between retinal imaging and MRI scan between the two groups. (Table 1)

**Table 1.**
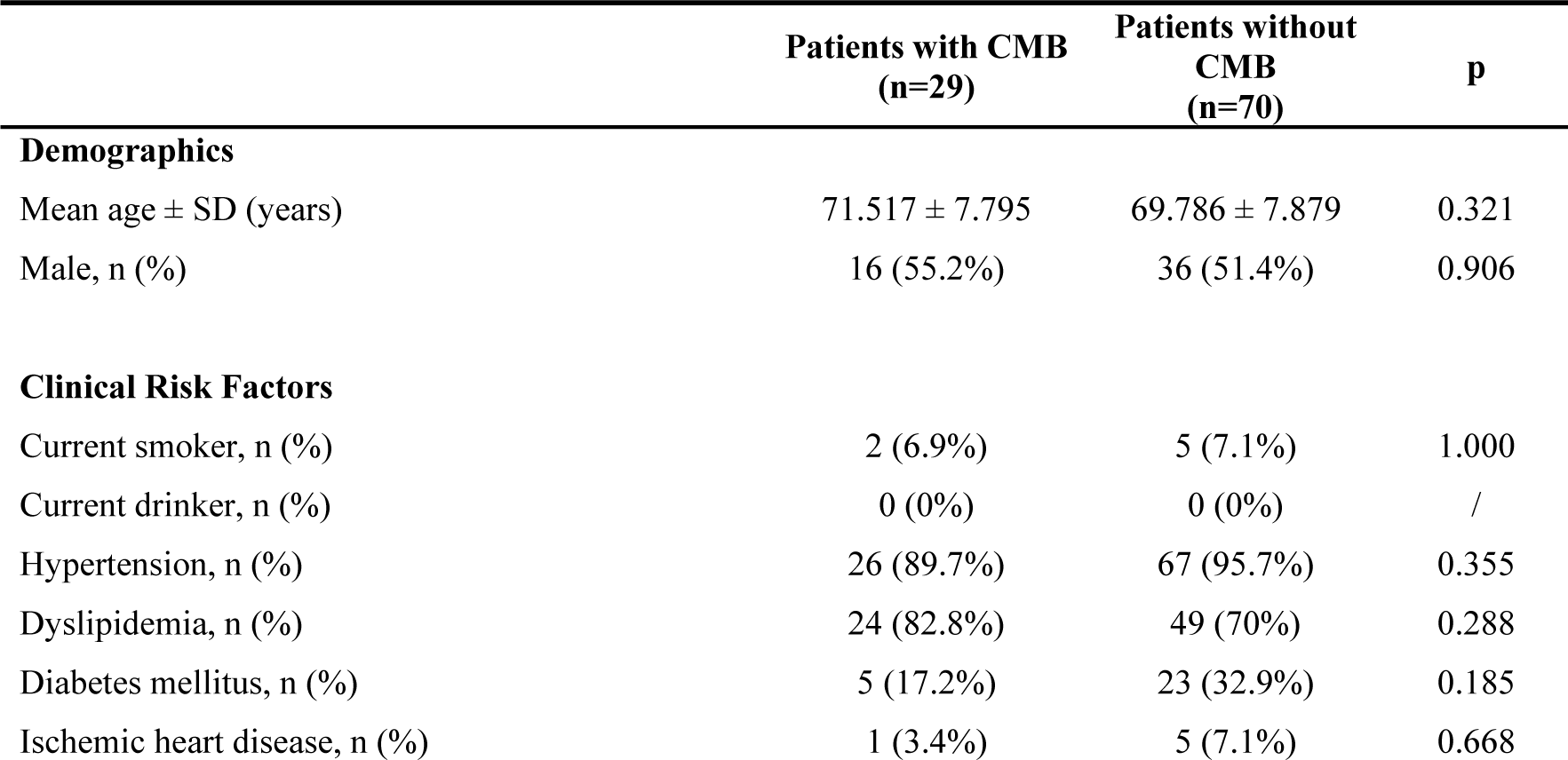
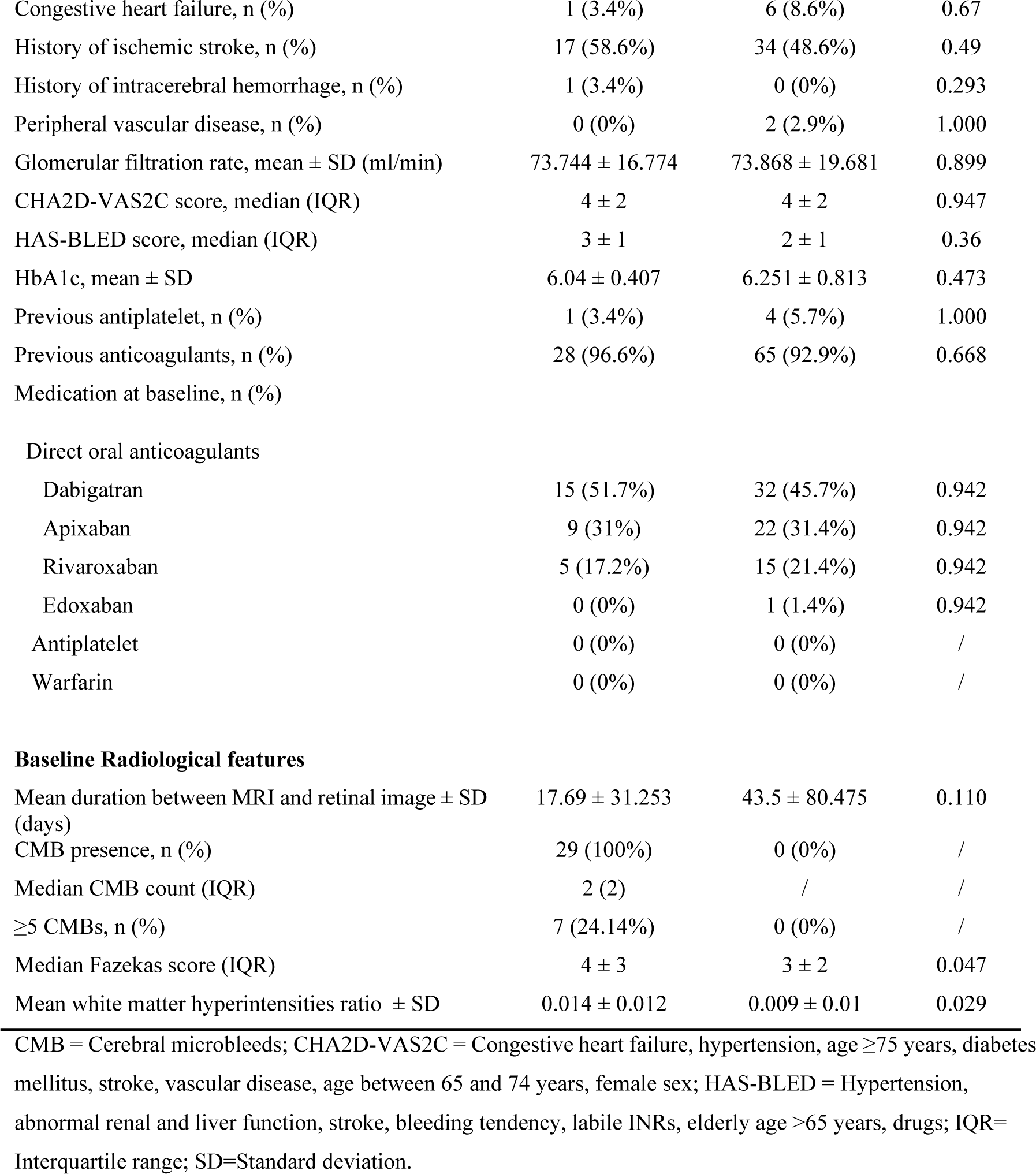
Characteristics of patients with atrial fibrillation, with and without cerebral *microbleeds*.

|  | Patients with CMB<br>(n=29) | Patients without<br>CMB<br>(n=70) | p |
| --- | --- | --- | --- |
| **Demographics** |  |  |  |
| Mean age ± SD (years) | $71.517 \pm 7.795$ | $69.786 \pm 7.879$ | 0.321 |
| Male, n (%) | 16 (55.2%) | 36 (51.4%) | 0.906 |
| **Clinical Risk Factors** |  |  |  |
| Current smoker, n (%) | 2 (6.9%) | 5 (7.1%) | 1.000 |
| Current drinker, n (%) | 0 (0%) | 0 (0%) | / |
| Hypertension, n (%) | 26 (89.7%) | 67 (95.7%) | 0.355 |
| Dyslipidemia, n (%) | 24 (82.8%) | 49 (70%) | 0.288 |
| Diabetes mellitus, n (%) | 5 (17.2%) | 23 (32.9%) | 0.185 |
| Ischemic heart disease, n (%) | 1 (3.4%) | 5 (7.1%) | 0.668 |
| Congestive heart failure, n (%) | 1 (3.4%) | 6 (8.6%) | 0.67 |
| History of ischemic stroke, n (%) | 17 (58.6%) | 34 (48.6%) | 0.49 |
| History of intracerebral hemorrhage, n (%) | 1 (3.4%) | 0 (0%) | 0.293 |
| Peripheral vascular disease, n (%) | 0 (0%) | 2 (2.9%) | 1.000 |
| Glomerular filtration rate, mean ± SD (ml/min) | 73.744 ± 16.774 | 73.868 ± 19.681 | 0.899 |
| CHA2D-VAS2C score, median (IQR) | 4 ± 2 | 4 ± 2 | 0.947 |
| HAS-BLED score, median (IQR) | 3 ± 1 | 2 ± 1 | 0.36 |
| HbA1c, mean ± SD | 6.04 ± 0.407 | 6.251 ± 0.813 | 0.473 |
| Previous antiplatelet, n (%) | 1 (3.4%) | 4 (5.7%) | 1.000 |
| Previous anticoagulants, n (%) | 28 (96.6%) | 65 (92.9%) | 0.668 |
| Medication at baseline, n (%) |  |  |  |
| Direct oral anticoagulants |  |  |  |
| Dabigatran | 15 (51.7%) | 32 (45.7%) | 0.942 |
| Apixaban | 9 (31%) | 22 (31.4%) | 0.942 |
| Rivaroxaban | 5 (17.2%) | 15 (21.4%) | 0.942 |
| Edoxaban | 0 (0%) | 1 (1.4%) | 0.942 |
| Antiplatelet | 0 (0%) | 0 (0%) | / |
| Warfarin | 0 (0%) | 0 (0%) | / |
| **Baseline Radiological features** |  |  |  |
| Mean duration between MRI and retinal image ± SD (days) | 17.69 ± 31.253 | 43.5 ± 80.475 | 0.110 |
| CMB presence, n (%) | 29 (100%) | 0 (0%) | / |
| Median CMB count (IQR) | 2 (2) | / | / |
| $\geq 5$ CMBs, n (%) | 7 (24.14%) | 0 (0%) | / |
| Median Fazekas score (IQR) | 4 ± 3 | 3 ± 2 | 0.047 |
| Mean white matter hyperintensities ratio ± SD | 0.014 ± 0.012 | 0.009 ± 0.01 | 0.029 |

CMB = Cerebral microbleeds; CHA2D-VAS2C = Congestive heart failure, hypertension, age $\geq 75$ years, diabetes mellitus, stroke, vascular disease, age between 65 and 74 years, female sex; HAS-BLED = Hypertension, abnormal renal and liver function, stroke, bleeding tendency, labile INRs, elderly age $>65$ years, drugs; IQR= Interquartile range; SD=Standard deviation.

### OCT-A Metrics of Patients with and without CMB

Compared to patients without CMB, patients with CMB had significantly larger mean FAZ at SCP, lower mean VD at SCP and larger vessel caliber with higher mean VDI at SCP, DCP and disc center. (Table 2) A trend of larger mean FAZ and lower mean VD in DCP were also noted in patients with CMB compared to patients without CMB.

**Table 2.** OCT-A metrics of patients with atrial fibrillation, with and without cerebral microbleeds.

|  | **Patients with**<br>**CMB**<br>**(n=29)** | **Patients without**<br>**CMB**<br>**(n=70)** | **p** |
| --- | --- | --- | --- |
| **Macular-centered OCT-A metrics at SCP** | **Mean ± SD** | **Mean ± SD** |  |
| FAZ area (mm<sup>2</sup>) | 0.579 ± 0.204 | 0.387 ± 0.126 | <0.001 |
| VD (%) | 76.5 ± 6.2 | 82.1 ± 6.7 | <0.001 |
| FAZ ratio circularity | 0.647 ± 0.089 | 0.62 ± 0.093 | 0.186 |
| Fractal dimension | 1.681 ± 0.010 | 1.682 ± 0.009 | 0.751 |
| VDI | 0.014 ± 0.001 | 0.013 ± 0.001 | <0.001 |
| SCP Image Quality Index | 66.241 ± 6.022 | 66.643 ± 4.848 | 0.751 |
| **Macular-centered OCT-A metrics at DCP** | **Mean ± SD** | **Mean ± SD** |  |
| FAZ area (mm<sup>2</sup>) | 1.530 ± 0.583 | 1.315 ± 0.541 | 0.059 |
| VD (%) | 38.5 ± 3.1 | 37.2 ± 2.8 | 0.057 |
| FAZ ratio circularity | 0.385 ± 0.108 | 0.366 ± 0.093 | 0.400 |
| Fractal dimension | 1.690 ± 0.013 | 1.691 ± 0.015 | 0.838 |
| VDI | 0.013 ± 0.001 | 0.011 ± 0.001 | <0.001 |
| DCP Image Quality Index | 66.241 ± 6.022 | 66.643 ± 4.848 | 0.751 |
| **Disc-centered OCT-A metrics, 3x3mm** | **Mean ± SD** | **Mean ± SD** |  |
| VDI | 0.017 ± 0.002 | 0.013 ± 0.001 | <0.001 |
| Fractal dimension | 1.633 ± 0.036 | 1.636 ± 0.030 | 0.562 |
| Mean Disc Image Quality Index | 69.931 ± 3.863 | 68.443 ± 4.702 | 0.107 |

CMB=cerebral microbleeds; OCT-A=Optical coherence tomography-angiogram; SCP=Superficial capillary plexus; FAZ=Foveal avascular zone; VD= vessel density; VDI=Vessel diameter index; SD=Standard deviation; DCP=Deep capillary plexus.

In multivariable logistic regression models, a larger FAZ area at SCP (OR 3.643 per SD increase, 95% CI 1.944 – 6.827), higher VDI across SCP (OR 4.733 per SD increase, 95% CI 2.414 – 9.281), DCP (OR 9.924 per SD increase, 95% CI 3.620 – 27.204) and disc center (OR 36.826 per SD increase, 95% C 7.990 – 169.739), as well as lower VD at SCP (OR 2.589 per SD decrease, 95% CI 1.455 – 4.609) were significantly associated with presence of CMB after adjusting for age, history of diabetes mellitus, white matter hyperintensities ratio, image quality index, duration between retinal imaging and MRI scan. (Figure 2)

**Figure 2.**
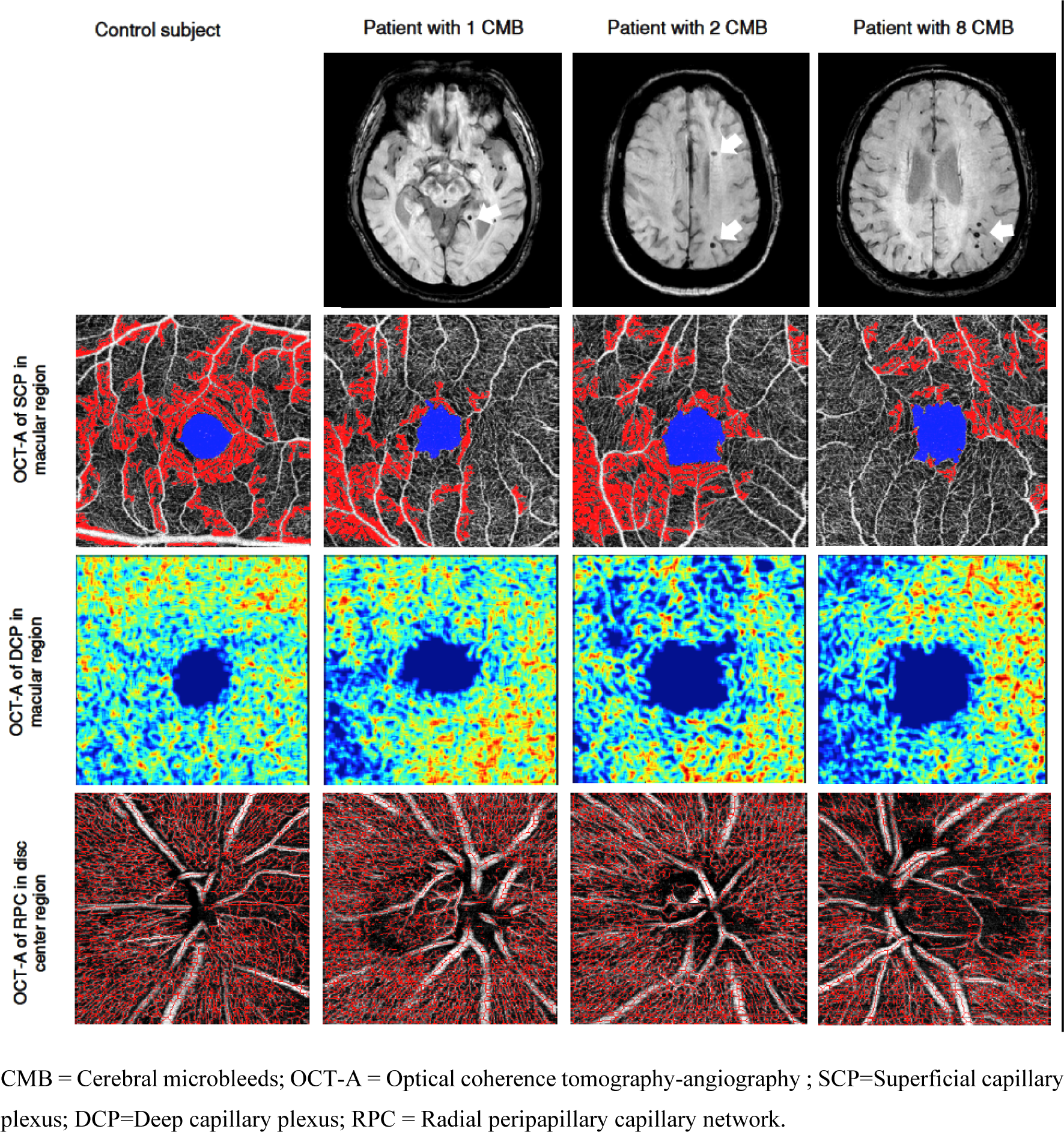
MRI and OCT-A metrics of control subject and patients with atrial fibrillation and CMB (arrows). Compared to control subject, patients with CMB had larger foveal avascular zone (blue area at SCP in the macular region), lower vessel density (larger blue area at DCP in the macular region) and larger vessel calibre (red area at RPC in the disc center).

With quasi-poisson regression, lager FAZ area at SCP (RR 1.740 per SD increase; 95% CI, 1.186 – 2.554) and higher VDI across SCP (RR 2.639 per SD increase; 95% CI, 1.448 – 4.810), DCP (RR 2.184 per SD increase; 95% CI, 1.405 – 3.396) and disc center (RR 1.878 per SD increase; 95% CI, 1.185 – 2.977) were associated with higher CMB count after adjustment for same confounding factors above. (Figure 2)

### OCT-A Metrics of Patients with AF vs Control Group

Compared to the 60 control subjects, the 99 patients with AF were significantly older (mean age 70.293±7.855 in AF vs 66.717±4.54 in control group, p =0.003) with more male patients (52.5% with AF vs 30% in control group, p=0.009). The abnormal retinal vascular parameters associated with presence of CMB (i.e. larger FAZ area and higher VDI in SCP, DCP and disc center) were also consistently observed in patients with AF compared to the control group. (Table 3, Figure 2)

**Table 3.** OCT-A metrics of patients with atrial fibrillation and control subjects.

|  | **Patients with AF**<br>**(n=99)** | **Control subjects**<br>**(n=60)** | **p** |
| --- | --- | --- | --- |
| **Demographics** |  |  |  |
| Mean age ± SD (years) | 70.293 ± 7.855 | 66.717 ± 4.540 | 0.003 |
| Male, n (%) | 52 (52.5%) | 18 (30%) | 0.009 |
| **Macular-centered OCT-A metrics at SCP** |  |  |  |
|  | **Mean ± SD** | **Mean ± SD** |  |
| FAZ area (mm<sup>2</sup>) | 0.443 ± 0.176 | 0.340 ± 0.110 | <0.001 |
| VDI | 0.013 ± 0.001 | 0.013 ± 0.000 | 0.001 |
| **Macular-centered OCT-A metrics at DCP** |  |  |  |
|  | **Mean ± SD** | **Mean ± SD** |  |
| VDI | 0.012 ± 0.001 | 0.011 ± 0.000 | 0.006 |
| **Disc-centered OCT-A metrics, 3x3mm** |  |  |  |
|  | **Mean ± SD** | **Mean ± SD** |  |
| VDI | 0.014 ± 0.002 | 0.013 ± 0.000 | <0.001 |

AF=atrial fibrillation; OCT-A= Optical coherence tomography-angiography; CMB=cerebral microbleeds; SCP=Superficial capillary plexus; FAZ=Foveal avascular zone; VD= vessel density; VDI=Vessel diameter index; DCP=Deep capillary plexus; SD=Standard deviation.

### OTCA Metrics of Patients with and without New CMB

Among the 99 patients with AF recruited into this study, 41 patients had previous MRI brain performed in the IPAAC registry. The median duration between the previous and index MRI was 35.09 months (IQR 33.544, 38.571). Five patients had new CMB on follow-up MRI, which were the index MRI for the current study. Patients with new CMB had wider vessel caliber with higher mean VDI in DCP and at the disc center. (Table 4)

**Table 4.** OCT-A metrics of 41 patients with atrial fibrillation who had previous MRI brain performed.

|  | Patients AF and new<br>CMB on follow-up<br>MRI<br>(n=5) | Patients with AF<br>without new CMB on<br>follow-up MRI<br>(n=36) | p |
| --- | --- | --- | --- |
| **Macular-centered OCT-A metrics at SCP** |  |  |  |
| FAZ area (mm<sup>2</sup>) | 0.524 ± 0.069 | 0.440 ± 0.170 | 0.066 |
| VD (%) | 79.2 ± 4.0 | 79.9 ± 8.0 | 0.755 |
| FAZ ratio circularity | 0.623 ± 0.057 | 0.628 ± 0.086 | 0.870 |
| Fractal dimension | 1.690 ± 0.007 | 1.680 ± 0.009 | 0.024 |
| VDI | 0.014 ± 0.001 | 0.013 ± 0.001 | 0.148 |
| SCP Image Quality Index | 68.600 ± 5.459 | 65.889 ± 5.290 | 0.343 |
| **Macular-centered OCT-A metrics at DCP** |  |  |  |
| FAZ area (mm<sup>2</sup>) | 1.650 ± 0.577 | 1.43 ± 0.624 | 0.461 |
| VD (%) | 35.9 ± 2.5 | 37.1 ± 2.9 | 0.356 |
| FAZ ratio circularity | 0.327 ± 0.091 | 0.377 ± 0.098 | 0.296 |
| Fractal dimension | 1.685 ± 0.014 | 1.688 ± 0.014 | 0.652 |
| VDI whole image | 0.013 ± 0.001 | 0.012 ± 0.001 | 0.012 |
| Mean DCP Image Quality Index | 68.600 ± 5.459 | 65.889 ± 5.290 | 0.343 |
| **Disc-centered OCTA metrics, 3x3mm** |  |  |  |
| VDI | 0.017 ± 0.002 | 0.014 ± 0.001 | <0.001 |
| Fractal dimension | 1.626 ± 0.031 | 1.632 ± 0.027 | 0.700 |
| Mean Disc Image Quality Index | 68.400 ± 5.595 | 68.972 ± 4.890 | 0.837 |

AF=atrial fibrillation; CMB=cerebral microbleeds; SCP=Superficial capillary plexus; FAZ=Foveal avascular zone; VD= vessel density; VDI=Vessel diameter index; DCP=Deep capillary plexus; SD=Standard deviation; VD = Vessel density.

## Discussion

In patients with AF who underwent both MRI brain and OCT-A, patients with CMB on MRI brain were more likely to have alterations in retinal capillary network compared to those without CMB. A larger FAZ and lower VD in SCP, as well as wider vessel caliber with higher VDI across SCP and DCP at macular region, along with the radial peripapillary capillary network at disc center, were associated with the presence and burden of CMB. In addition, higher VDI at DCP and disc center were also associated with development of new CMB. Compared with healthy control, FAZ and VDI across SCP, DCP and disc area remained consistently different between patients with AF and control group.

To our knowledge, this is the first study evaluating the associations of OCT-A metrics changes with CMB in patients with AF. Lange et al have reported reduced flow density measured by OCT-A of 44 patients with AF compared to control group, which was possibly related to reduced cardiac output with AF.(21) In our study, we studied extensively the integrity of the retinal capillary network (including FAZ, vessel caliber, vessel density and branching) using OCT-A in patients with AF and investigated their changes associated with presence of CMB on MRI brain. As white matter hyperintensities, which commonly co-exist with CMB, can be associated with many other conditions including aging, dementia, previous ischaemic stroke and diabetes mellitus etc., we adjusted for this confounding factor meticulously by semiautomated segmentation method taking into account the parenchymal brain volume. Among all the OCT-A metrics found to be associated with CMB, increase in FAZ at SCP as well as VDI across SCP, DCP at macular region and the radial peripapillary capillary network at center showed most consistent associations with presence and burden of CMB. These metrics were also significantly different between patients with AF and control group. On the other hand, patients with AF and CMB also had trend of larger FAZ and higher VDI at DCP compared to those without CMB. It remains possible that these may also be potential parameters associated with CMB if the study was performed with a larger sample size. While the exact mechanism underlying these vascular changes are uncertain, we hypothesize that it could be related to capillary drop-out and secondary vasodilatation at the capillary layer. In our study, we have excluded 20 subjects, mostly patients with AF, with suboptimal retinal image quality. As OCT-A requires patient’s cooperation to fixate and follow commands well for the imaging, it is more suitable for patients with AF without significant residual neurological deficit from stroke and those who had not experienced stroke. The later, particularly those who are young with very few risk factors for stroke, are most in need of risk stratification tool for intracerebral hemorrhage, as their risk of ischaemic stroke may be relatively low, that the risk of intracerebral hemorrhage associate with anticoagulation is of major concern to them during decision-making. OCT-A may offer prognostic information when MRI brain is otherwise not indicated.

The major challenge of the study was the disruption of study schedule by the COVID-19 pandemic and social unrest in Hong Kong, that more than one-third of the patients had a duration between retinal imaging and MRI scan for more than 2 weeks. Although this duration was not statistically different between patients with and without CMB, we have adjusted for this factor in all our analyses to minimise any influence incurred. Other limitations include i) patients with poor mobility and use of other anti-thrombotic agents, who have higher risk of vascular events, were not included in this study. Hence, the results cannot be applied to this spectrum of patients; ii) the proportion of patients with CMB in patients with AF were lower than that assumed in the sample size estimation. However, we also had a higher proportion of patients with ≥5 CMB than expected, which helped provide a reasonable sample size of patients with moderate CMB burden for this study. Further studies with larger sample size will be needed to replicate the findings; iii) our control group was significantly younger than patients with AF as it was difficult to identify subjects above 70 years old without significant medical history. Hence, we have adjusted for age in all our analyses; iv) only Chinese patients are included in this study, further studies including other ethnic groups are needed.

The strength of this study include i) patients recruited in this study were highly homogeneous that all were receiving direct oral anticoagulants, which have been shown not to affect the development of CMB;(11) ii) background of patients with and without CMB was highly comparable, minimising confounders in the analysis; iii) A control group was available to ensure the relevance of findings when used in the community setting; iv) with robust adjustment for confounding factors and image quality, we were able to achieve consistent findings as mentioned despite a relatively small sample size

## Conclusions

Due to aging population, there has been progressive increase in prevalence of AF, affecting up to 10-17% among those aged above 80 years old. The number is projected to double by 2060, posing significant healthcare burden.(3) Current efforts are dedicated to preventing the occurrence of stroke secondary to AF with various strategies which have been continuously evolving. Developing a non-invasive tool with OCT-A, complementary to MRI brain, to assess risk of anticoagulant-associated intracerebral hemorrhage may help provide further prognostic information for patients with AF in the community, particularly for those who have otherwise no indication or have contraindications for MRI brain.

This pilot study provides preliminary data on how the retinal capillary network changes in the presence of bleeding-prone microangiopathy with CMB among patients with AF on anticoagulant. Larger FAZ at SCP as well as higher VDI across SCP, DCP and disc center were consistently associated with presence and burden of CMB. The findings suggest the potential role of OCT-A to assist risk stratification for patients with AF who are require anticoagulation in the future. Further studies focusing on FAZ and VDI with larger sample size with different ethnic groups should be performed to determine the cut-off values which best identify patients with multiple CMB who have significantly increased risk of anticoagulant-associated intracerebral.

## Data Availability

Annqonymized data of this study will be available from Corresponding Author upon justified request.

## Acknowledgements

We acknowledge the contribution by Mr Trevor Lau, Ms Haley Mak and Ms Annie Ling in retinal imaging.

